# Replacement of the Alpha variant of SARS-CoV-2 by the Delta variant in Lebanon between April and June 2021

**DOI:** 10.1101/2021.08.10.21261847

**Authors:** Georgi Merhi, Alexander J. Trotter, Leonardo de Oliveira Martins, Jad Koweyes, Thanh Le-Viet, Hala Abou Naja, Mona Al Buaini, Sophie J. Prosolek, Nabil-Fareed Alikhan, Martin Lott, Tatiana Tohmeh, Bassam Badran, Orla J. Jupp, Sarah Gardner, Matthew W. Felgate, Kate A. Makin, Janine M. Wilkinson, Rachael Stanley, Abdul K. Sesay, Mark A. Webber, Rose K. Davidson, Nada Ghosn, Mark Pallen, Hamad Hasan, Andrew J. Page, Sima Tokajian

## Abstract

**Background:** The COVID-19 pandemic continues to expand globally, with case numbers rising in many areas of the world, including the Eastern Mediterranean Region. Lebanon experienced its largest wave of COVID-19 infections from January to April 2021. Limited genomic surveillance was undertaken, with just twenty six SARS-CoV-2 genomes available for this period, nine of which were from travellers from Lebanon detected by other countries. Additional genome sequencing is thus needed to allow surveillance of variants in circulation.

**Methods:** Nine hundred and five SARS-CoV-2 genomes were sequenced using the ARTIC protocol. The genomes were derived from SARS-CoV-2-positive samples, selected retrospectively from the sentinel COVID-19 surveillance network, to capture diversity of location, sampling time, gender, nationality and age.

**Results:** Although sixteen PANGO lineages were circulating in Lebanon in January 2021, by February there were just four, with the Alpha variant accounting for 97% of samples. In the following two months, all samples contained the Alpha variant. However, this had changed dramatically by June and July, when all samples belonged to the Delta variant.

**Discussion:** This study provides a ten-fold increase in the number of SARS-CoV-2 genomes available from Lebanon. The Alpha variant, first detected in the UK, rapidly swept through Lebanon, causing the country’s largest wave to date, which peaked in January 2021. The Alpha variant was introduced to Lebanon multiple times despite travel restrictions, but the source of these introductions remains uncertain. The Delta variant was detected in Gambia in travellers from Lebanon in mid-May, suggesting community transmission in Lebanon several weeks before this variant was detected in the country. Prospective sequencing in June/July 2021 showed that the Delta variant had completely replaced the Alpha variant in under six weeks.

## Introduction

Since the first report from Wuhan, China in December 2019 (Huang et al. 2020), Covid-19 has spread globally. The causative agent, severe acute respiratory syndrome-related coronavirus 2 (SARS-CoV-2), has caused by July 31 >200 million cases and > four million deaths (Dong, Du, and Gardner 2020) worldwide and >562,527 cases and 7,909 deaths In Lebanon (https://www.moph.gov.lb/en/). After a period of relative stasis, since late 2020, the evolution of SARS-CoV-2 has been linked to Variants of Concern (VOCs), characterized by increased epidemic potential (Harvey et al. 2021) and distinctive sets of mutations on genome sequencing. VOCs have been assigned letters of the Greek alphabet in the WHO naming scheme (Konings et al). Of these, the Alpha variant (PANGO lineage B.1.1.7) (Volz et al.2021; Davies et al.2021) was first detected in the United Kingdom in September 2020, while the Delta variant (PANGO lineage B.1.617.2) was first detected in India in October 2020 (WHO 2021).

The first case of COVID-19 was reported in Lebanon on February 21, 2020. From March to July 2020, the number of cases remained low, with less than a hundred cases daily. Subsequently, case numbers increased as a result of:

1. The reopening of Beirut airport to international travellers;
2. An explosion at the Beirut Port on 4 August—one of the largest non-nuclear explosions ever recorded (Rigby et al. 2020)—displacing much of the population of the city;
3. the easing of the lockdown restrictions, leading by December 2020 to record highs (over 1,000) in the number of COVID-19 cases per day (Dong, Du, and Gardner 2020).

The first case of the Alpha variant in Lebanon was detected on 22 December in a traveller arriving from the UK. This was followed by a surge in cases, triggering increased containment measures and a vaccination campaign. As a result, by early June 2021 (Koweyes et al.2021) rates had returned to low levels. A subsequent loosening of restrictions led to the return of many Lebanese expatriates and by the end of June, case numbers had increased.

Sequencing of SAR-CoV-2 genomes from Lebanese samples has proven challenging because of:

1. reductions in air traffic have caused disruptions to supply chains, particularly those including a cold chain
2. increased demand for consumables in high-income countries have led to marked and prolonged shortages in low- and middle-income countries
3. the high per-sample costs of sequencing.

With the emergence of multiple variants of concern, the WHO has encouraged genomic surveillance worldwide. Rapid genome sequencing has been used widely in public health, in investigation of outbreaks (Page et al.2021), identification of novel variants of concern (Faria et al.2021; Tegally et al.2021, 351) and tracking the source and spread of variants (Mashe et al.2021). To provide a greater insight into the pandemic in Lebanon, we have retrospectively sequenced 905 SARS-CoV-2 samples collected from December 2020 to May 2021.

## Results

### Establishment of Alpha

The GISAID database houses thirty-nine SARS-COV-2 genome sequences that were generated before this study from samples taken in Lebanon or from individuals who had recently left the country (Table 1). These include eight samples related to travel outside of Lebanon and two genomes sequenced in Japan. Analyses of these genomes reveals that at least nine SARS-CoV-2 lineages were circulating in Lebanon up to late December 2020, with no single lineage dominating (Figure 4). The Alpha variant was first detected in Lebanon on 22 December 2020 in a traveller from the UK.

**Table 1:** PANGO lineages of GISAID data (2021-08-03) where Lebanon is the country of exposure, together with genomes sequenced in the present study

| Lineage | GISAID | This study |
| --- | --- | --- |
| B.1.1.7 | 23 | 782 |
| B.1.398 | 25 | 38 |
| B.1 | 8 | 18 |
| B.1.1 | 2 | 8 |
| B.1.36.1 |  | 7 |
| B.1.1.1 |  | 3 |
| B.1.177.86 |  | 2 |
| B.1.36 | 1 | 2 |
| B.1.438 | 1 | 2 |
| B.1.467 |  | 2 |
| B.1.1.315 | 1 | 1 |
| B.1.1.398 |  | 1 |
| B.1.160 |  | 1 |
| B.1.177 |  | 1 |
| B.1.177.53 |  | 1 |
| B.1.243 |  | 1 |
| B.1.258 |  | 1 |
| C.1 |  | 1 |
| L.3 | 1 | 1 |
| B.1.1.274 | 2 |  |
| B.1.1.487 | 1 |  |
| B.1.351 | 1 |  |
| B.1.36.35 | 1 |  |
| B.1.525 | 1 |  |
| B.1.617.2 | 8 |  |
| B.1.627 | 1 |  |

Among the genomes sequenced in this study, the first belonging to the Alpha variant was detected in a sample from the community taken on 22 December. However, by the second week in January 2021, the Alpha variant was present in five of Lebanon’s eight provinces and by the end of that month, had been detected in seven provinces (Supplementary Table 1), with the apparent absence in Akkar best explained by under-represented sampling. The Alpha variant remained the only VOC detected via community sequencing in Lebanon up until June 2021

There were no discernible links to gender, nationality or age during the establishment of the Alpha variant. Bayesian evolutionary analysis revealed some evidence of provincial clustering and subsequent seeding of other provinces, which is to be expected given the small size of the country (Figure 4 and Supplementary Figure 1). Phylogenetic analysis of alpha-variant genomes revealed considerable diversity, suggesting over forty independent importations (Figure 1).

**Figure 1:**
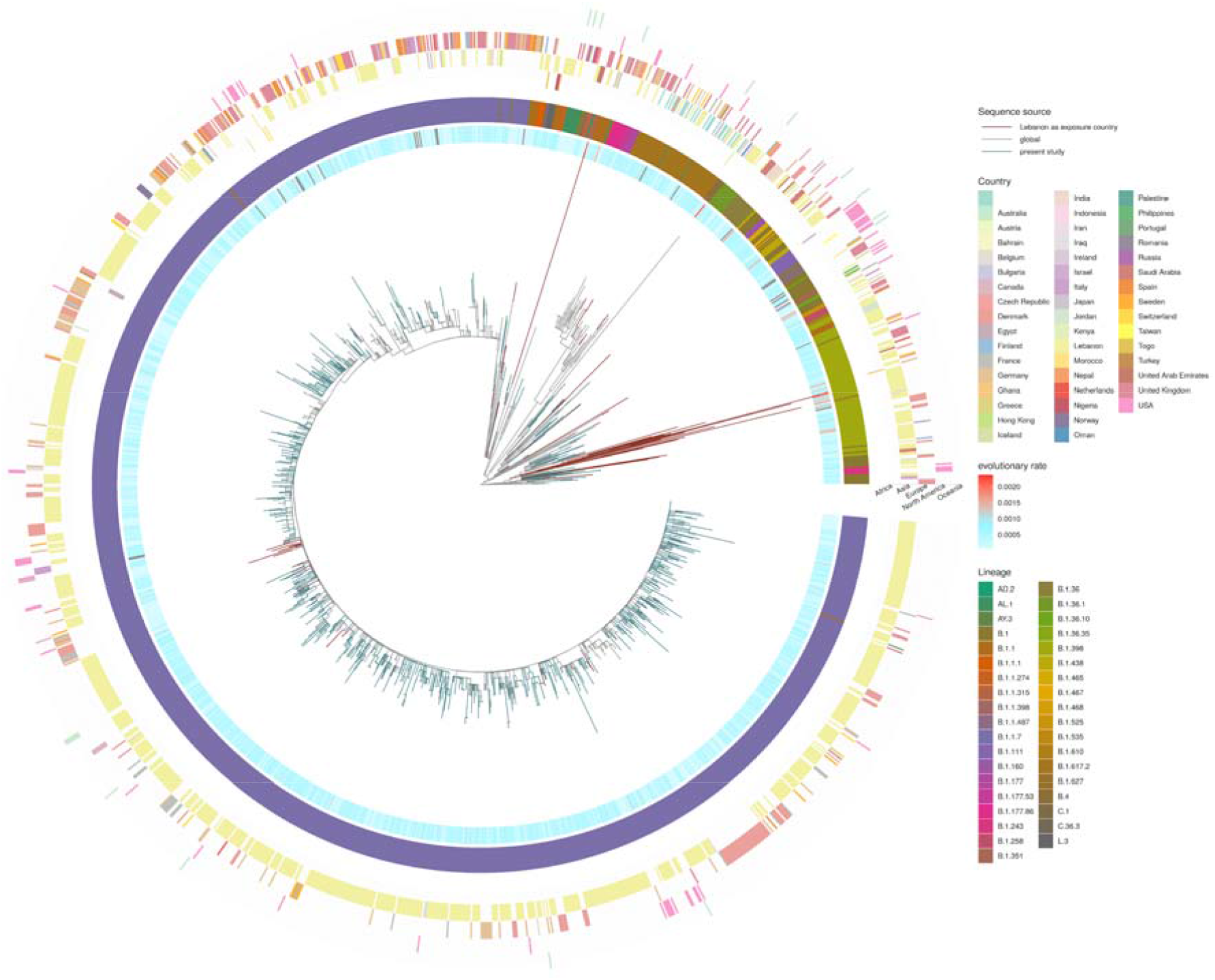
Phylogeny of B.1.1.7 including genomes from Lebanon. Samples sequenced for the current study (blue) together with GISAID sequences where Lebanon is identified as the source of exposure (brown) together with a few close sequences from abroad. The apparent evolutionary rate from a timetree analysis is shown, indicating sequences deviating from a molecular clock in red.

Comparisons with sequences in GISAID revealed that genomes identical to an Alpha variant from Lebanon were found in twenty-five countries (Figure 2). However, due to the slow mutation rate of SARS-CoV-2 and large variation in incubation time, it is not possible to unambiguously identify the transmission route from genomic data alone. Genomes from eleven countries have a perfect match to at least forty Lebanese genomes, with several European countries providing a larger number of matches than the UK, despite the UK providing for over 41% of the genomes from Europe and the location where the Alpha variant was first detected. This suggests that seeding of the Alpha variant into Lebanon probably occurred most often via intermediate countries rather than directly from the UK.

**Figure 2:**
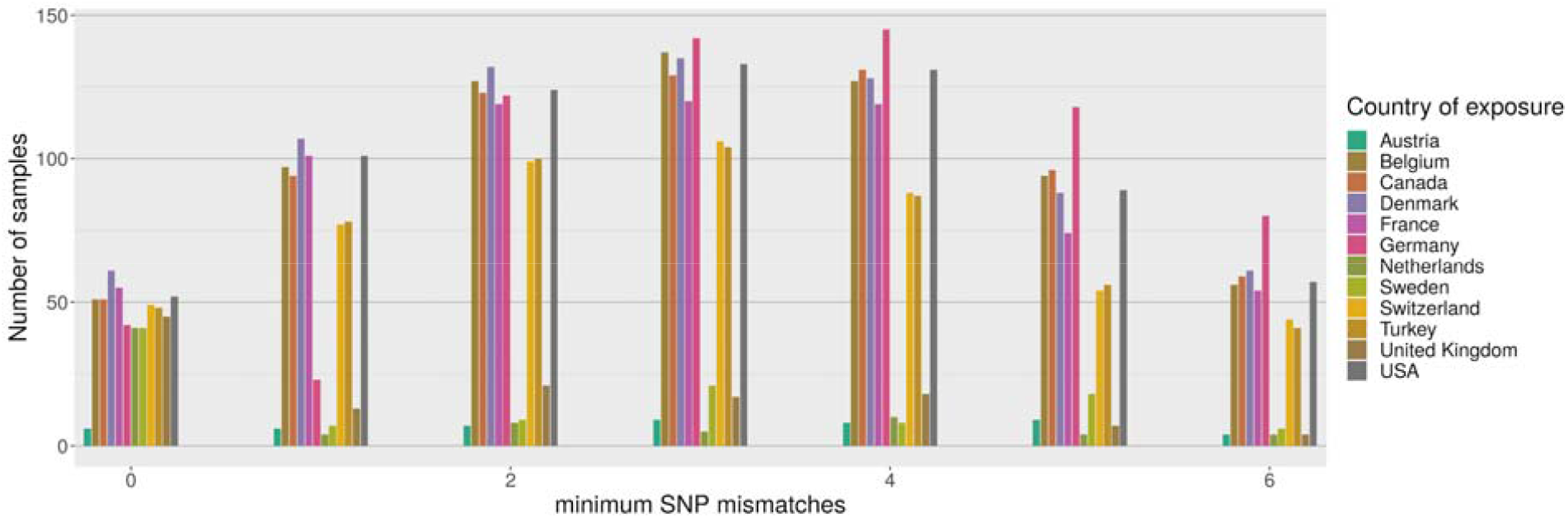
Distribution of best matches per country between local Alpha samples and global sequences from GISAID (2021-07-27). For each sample we find the 128 closest samples in GISAID and keep track of the best match per country. Only the most common countries are shown above, and the figure is truncated at 6 mismatches.

### Replacement with Delta

No genomes for samples collected in Lebanon are available for a 7 week period from 5/5/2021 to 24/6/2021, which corresponded to a period of very low case numbers. The first cases of Delta in The Gambia were detected in travellers from Lebanon (accession numbers: EPI_ISL_2820697, EPI_ISL_2820699) by genome sequencing at the MRC Unit The Gambia at the end of May 2021, indicating community transmission of Delta was already occurring in Lebanon. In that time there was a complete replacement of Alpha with Delta. A total of 28 samples were sequenced by LAU between 25/6/2021 and 3/7/2021, 100% of which were Delta (Supplementary Table 2). This rapid replacement of the dominant variant reflects the experience of many other countries worldwide, such as the UK, Denmark and the US (Shu and McCauley 2017).

**Figure 3:**
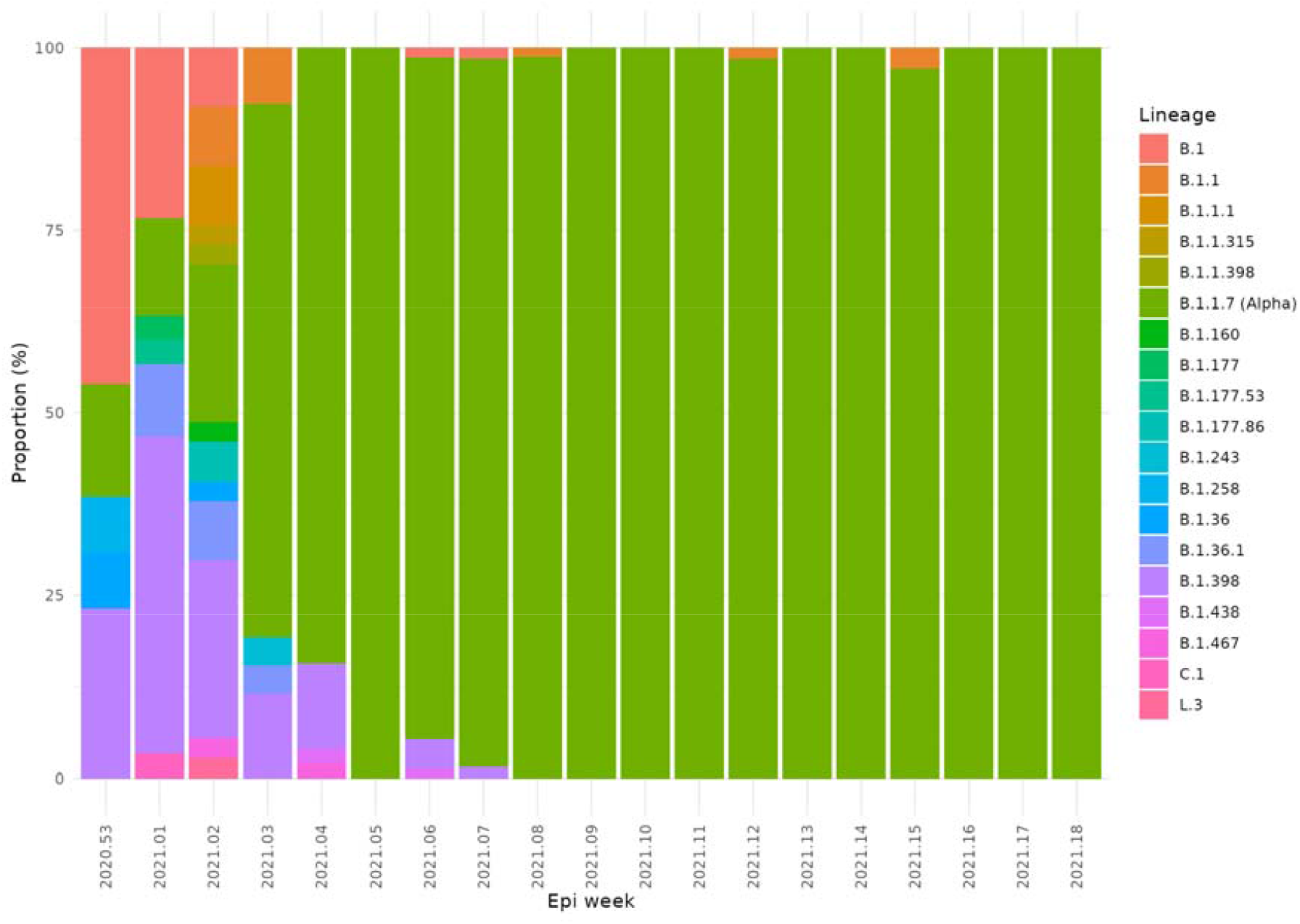
Change in prevalence of PANGO lineages between January to May 2021 in Lebanon showing the emergence and dominance of Alpha.

## Discussion

This study provides a 10 fold increase in the number of available SARS-CoV-2 genomes for Lebanon, providing insights into the dynamics of the pandemic. Lebanon shares a long land border with Syria, which is in the midst of a long running civil war, and has provided protection for millions of Syrian refugees. No public genomic surveillance has been undertaken in Syria (just 1 case detected by Japanese quarantine measures), so this dataset can help infer information about the pandemic in the wider region as infectious diseases know no borders.

The first COVID-19 cases in Lebanon starting February 2020 was dominated by lineage B.1. Phylogenetic analysis of whole genome sequences in a previous report (Feghali et al. 2021) and in this study confirmed multiple introduction scenarios due to international travel followed by community spread leading to strict lockdown measures. Genome sequencing facilities in LMIC remain scarce and are often associated with limited resources. In Lebanon, to the date of writing this manuscript, 1052 (905 from this study) SARS-CoV-2 genomes were deposited on GSAID, representing 2 of the overall sequenced genomes/1000 cases. Genome sequencing and phylogenetic analysis are key strategies to investigate and track the circulating viral lineages, identify routes of transmissions, and monitor and mitigate the spread of early introductions of variants of concerns.

Up until December 2020, no single variant completely dominated in Lebanon, although the B.1.398 lineage was a constant feature. This was in stark contrast to other countries which saw wholesale replacements of the wildtype with a single lineage. In Europe the B.1.177 lineage became the dominant lineage in Autumn 2020 after being widely dispersed from Spain in the summer of 2020, as it had gained the A222V mutation in the Spike protein which gave a fitness advantage.

The Alpha variant rapidly swept through Lebanon causing its largest wave to date, with cases peaking in January 2021. Following the emergence of Alpha in the UK, travellers from the UK to Lebanon were put through extra COVID-19 measures on top of the implemented ones. The first detected case of Alpha was on 22/12/2020 in a traveller from the UK. During the holiday period of December 2020, lockdown restrictions were released and were associated with an increase in the number of reported COVID-19 cases reaching 1,843/day on 31 December 2020 (Koweyes et al. 2021) and announcing the wave of January 2021. Later, the country went into a nationwide lockdown on January 7, 2021. Within this study, Alpha was detected in the community on the earliest collection date (29/12/2020) and 3 weeks later cases were detected in 5 out of 8 provinces, with Alpha accounting for 84% of cases (n=43/51) by the last week of January 2021.

However, it appears that travel guidelines and restrictions were insufficient to prevent transmission. Phylogenetic analysis showed that travel from the UK was not the primary driver for importation events of Alpha in Lebanon. Eleven countries have perfectly matching genomes to 40 Lebanese genomes indicating a recent common history. As can be seen in Figure 2 the UK is not the most closely associated country with Alpha genomes found in Lebanon, instead it is European countries including Belgium, Denmark, France, German and Switzerland, in addition to Turkey, Canada and USA. These countries have large Lebanese diaspora populations. Whilst directionality is difficult to attribute using genome sequencing alone, it indicates that the UK was not the primary driver of seeding events into Lebanon and that the travel restrictions placed on UK arrivals were effective.

The earliest traveller from Lebanon with Alpha was an individual who travelled to Singapore with a testing date of 16/1/2021. This would indicate that Alpha was already established in Lebanon at this point. This fits with the rapidly rising proportion of cases at this time from the genomes generated for this manuscript. The Alpha variant spread to every province in Lebanon. There is evidence of distinct clusters of Alpha establishing within particular provinces within Lebanon with onward seeding of other provinces. Exports from Lebanon to other countries were estimated to peak at the start of February (Figure 4 bottom panel). The public health measures which were introduced were sufficient to suppress the wild type lineages, however were insufficient to control Alpha which has increased transmissibility. This highlights the need for rapid prospective genomic surveillance to help inform public health measures.

**Figure 4:**
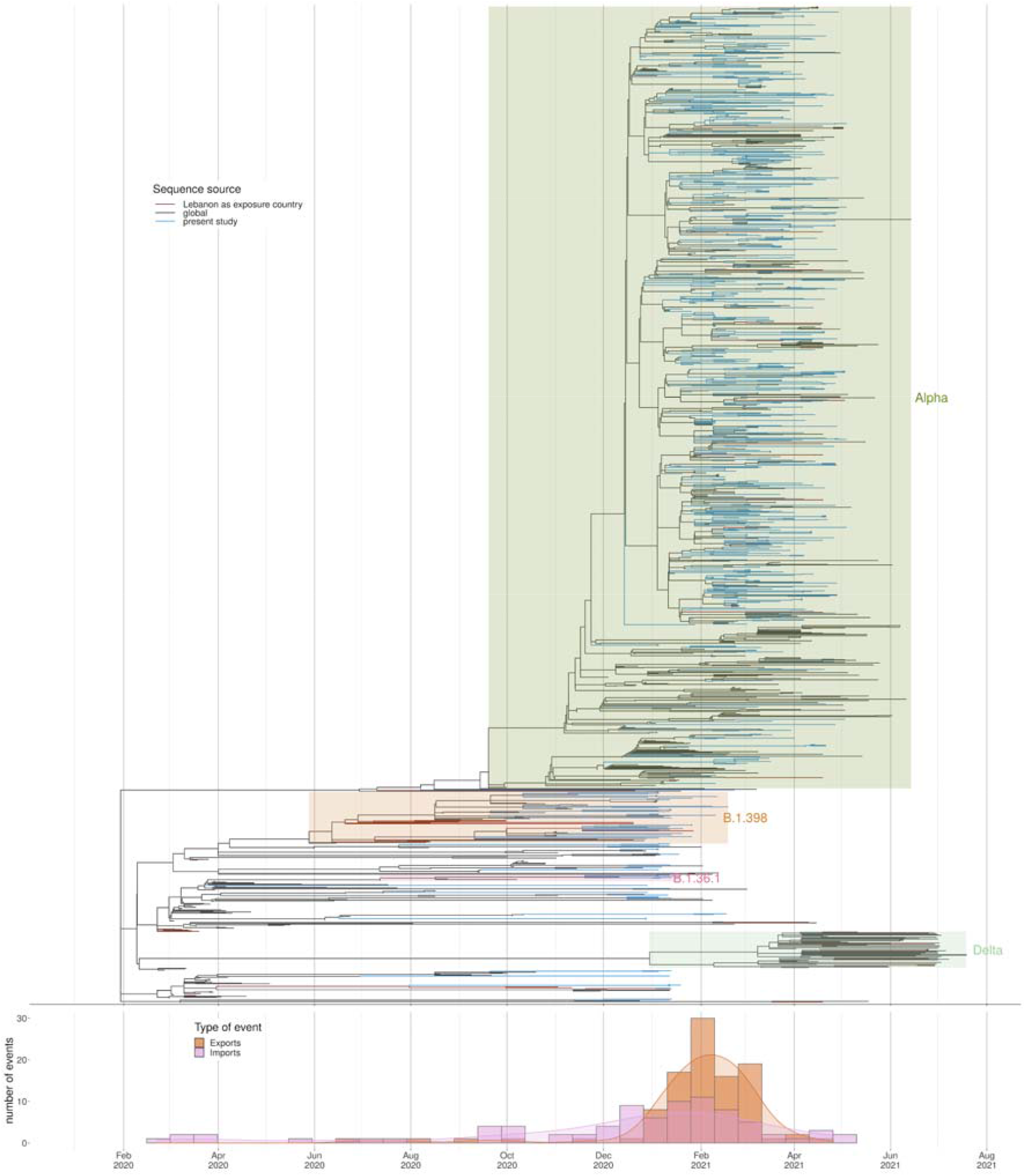
Divergence times estimation using all samples with complete data information, from all lineages. Below we have the estimated number of importations and exportations into/from Lebanon, based on the tree.

No other variants of concern were observed during this time period, up until the beginning of May 2021.

The Delta Variant of Concern was first identified in India and rapidly became the dominant variant in the UK in April 2021. The first indication that the Delta variant had reached Lebanon was in cases from Gambia with a travel history from Lebanon. These were the first identified cases of Delta in Gambia. This indicates that by the middle of May 2021 Delta was present in the community in Lebanon, a time when overall case numbers were low. Genomic surveillance started in Lebanon at the end of June 2020 and 100% of community cases were identified as Delta, confirming that the Delta variant was now widespread in the community. The importation events of Delta which seeded community transmission started silently many weeks before the official announcement of its discovery on the 2/7/2021. This highlights the need for ongoing rapid prospective genome sequencing of community cases in addition to testing at borders.

## Conclusion

Our results revealed how a variant could rapidly become dominant and that travel restrictions alone are insufficient to prevent transmission. We highlighted the importance of sequence-based surveillance in monitoring SARS-CoV-2 community transmission, identifying, and detecting emerging variants. Targeted travel restrictions did appear to be effective. Integrated genome sequencing plays a critical role in guiding interventions, containment strategies, and travel restrictions. Through WHO’s support, the recently launched national genomic surveillance initiative should be further expanded and supported to track the emergence, introduction, and spread of new SARS-CoV-2 variants.

## Data Availability

Assembled/consensus genomes are available from GISAID (Shu and McCauley 2017) subject to minimum quality control criteria. Raw reads are available from European Nucleotide Archive (ENA) in Bioproject PRJEB46859. Accession numbers, metrics and metadata are available for each sample in Supplementary Table 1.

## Ethics

The study protocol was ethically approved by the Lebanese American University, Institutional Review Board (LAU IRB) (reference LAU.SAS.ST2.19/May/2020). The Minister of Public Health for the Republic of Lebanon exempted the samples used for this study from the Nagoya protocol. The authors from the Ministry of Public Health and the National Influenza Center had access to patient identifiable information, all other authors had access to limited de-identified information on each patient.

## Acknowledgements

The authors wish to acknowledge the assistance of Nisrine Jammal, Lina Mroueh, Fadi Abdel Sater, Rawan Makki, Alissar Zaghlout, World Health Organization (WHO), United Nations High Commissioner for Refugees (UNHCR), United Nations Relief and Works Agency for Palestine Refugees in the Near East (UNRWA), Médecins Sans Frontières (MSF), Lebanese Municipalities, International Orthodox Christian Charities (IOCC), Islamic Health Society, Relief International, Amel Association, Primary Health Care Department at MOPH, Lebanese Red Cross, and the sentinel sites.

## Author contributions

Funding acquisition: AJP, ST, MP. Leadership and supervision: MW, RKD, MP, HH, AJP, ST. Metadata curation: TLV, NFA, ML, AKS, NG, HAN, MAB. Samples and logistics: MAB, AJT, SJP, TT, BB, OJJ, SG, MWF, KAM, JMW, RS, RKD, NG, HAN. Sequencing and analysis: GM, AJT, LdOM, JK, TLV, SJP, NFA, AJP. Visualization: LdOM, TLV.

## Supplementary

Supplementary Table 1: Spreadsheet of sample metadata and accession numbers.

Supplementary Table 2: GISAID acknowledgements.

## Methods

### Samples

For this study a total of 905 SARS-CoV-2 positive samples were collected from Lebanon, selected at random from clinical diagnostic samples, to provide a representative view of regions within Lebanon as shown in Supplementary Figure 1. The samples represented a broad range of ages and genders (Supplementary Figure 2) and were collected from a wide range of nationalities (Supplementary Figure 3). All cases were symptomatic at time of sample collection.

**Supplementary Figure 1:**
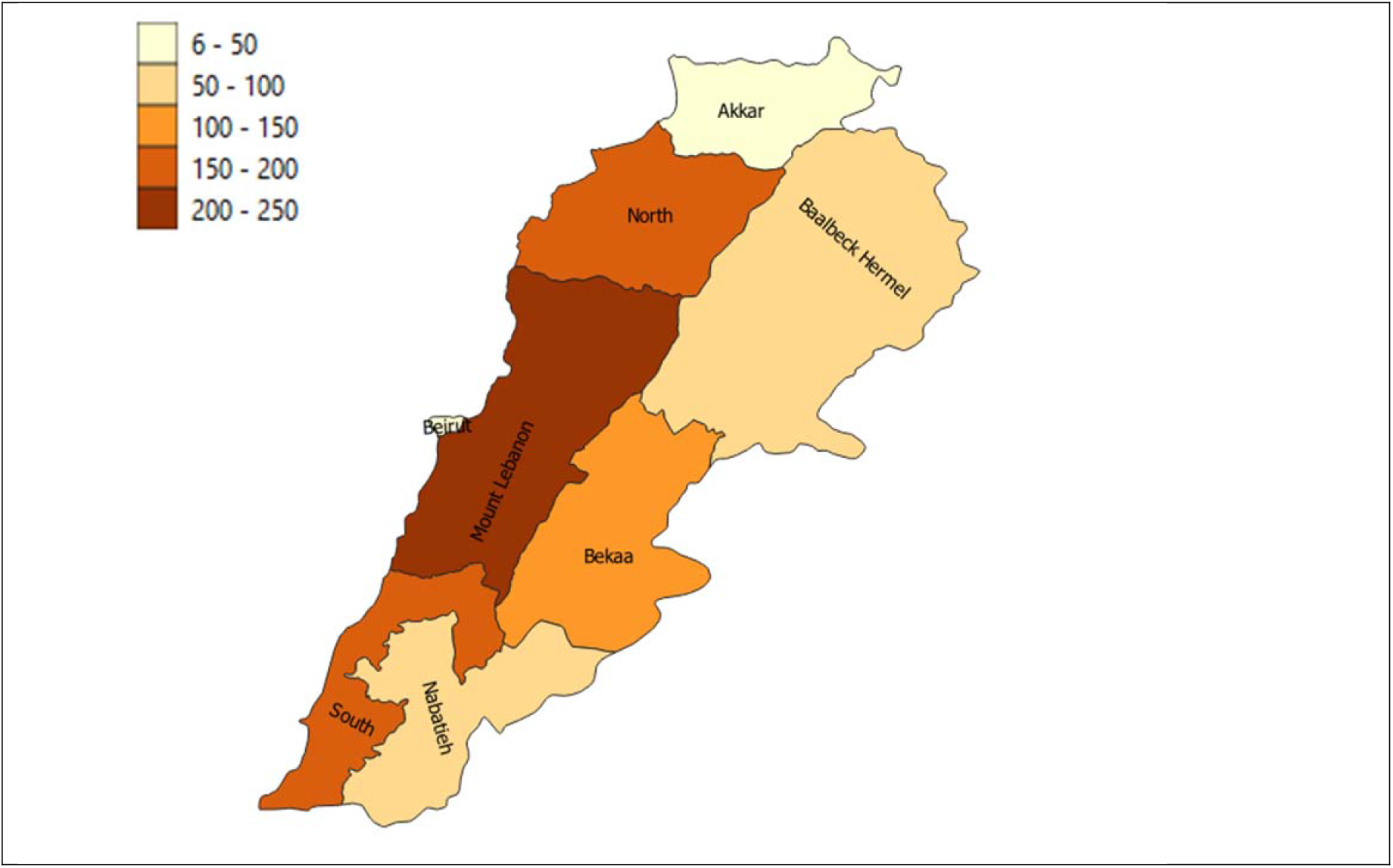
Distribution of sequenced cases in this study for each province.

**Supplementary Figure 2:**
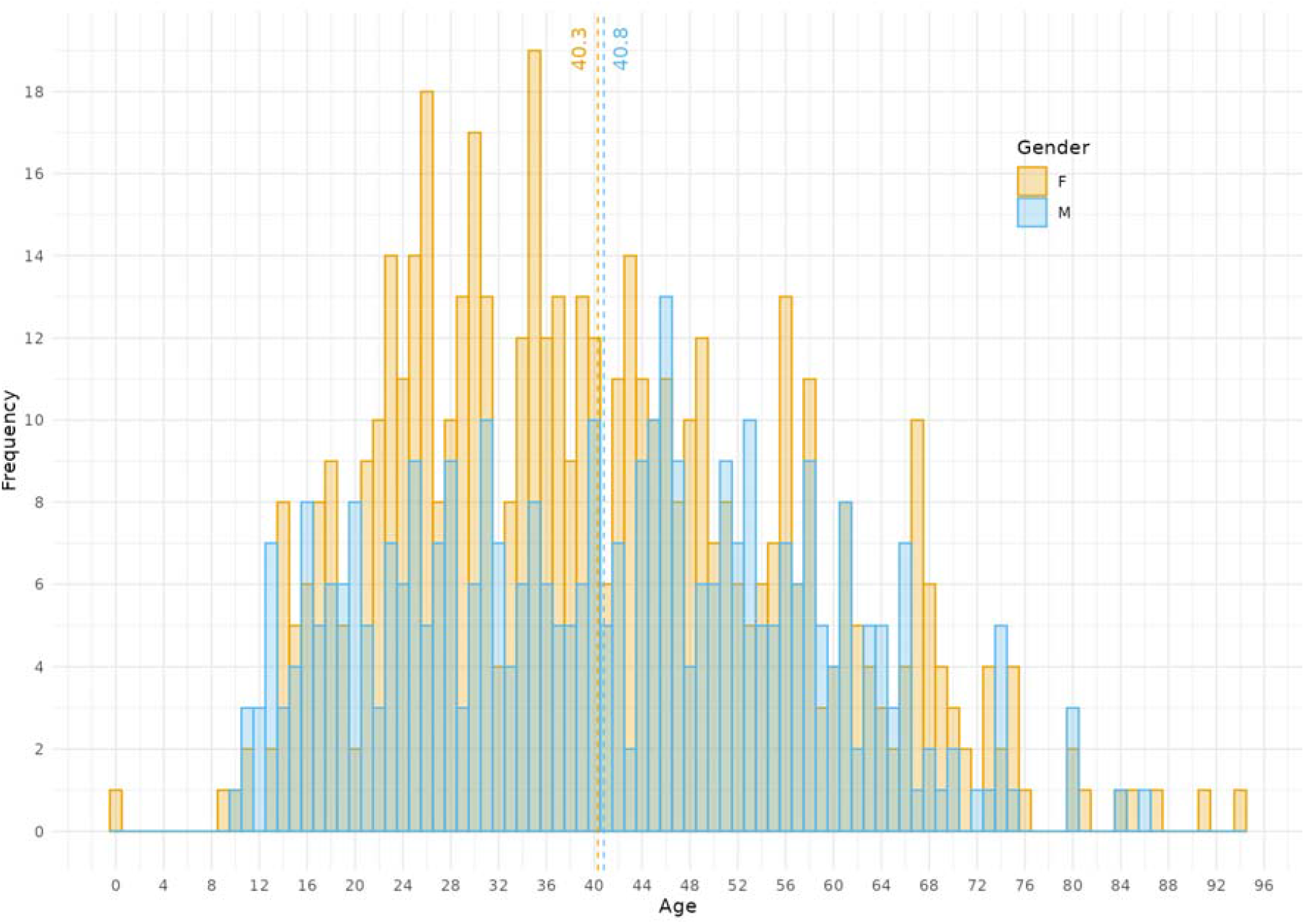
Demographics of samples from cases included in this study. Two cases with unknown gender were excluded. An age of zero corresponds to a baby under the age of 1. The mean age of the female cases was 40.3 years (± 16.5 SD) and the male cases was 40.8 years (± 16.8 SD).

**Supplementary Figure 3:**
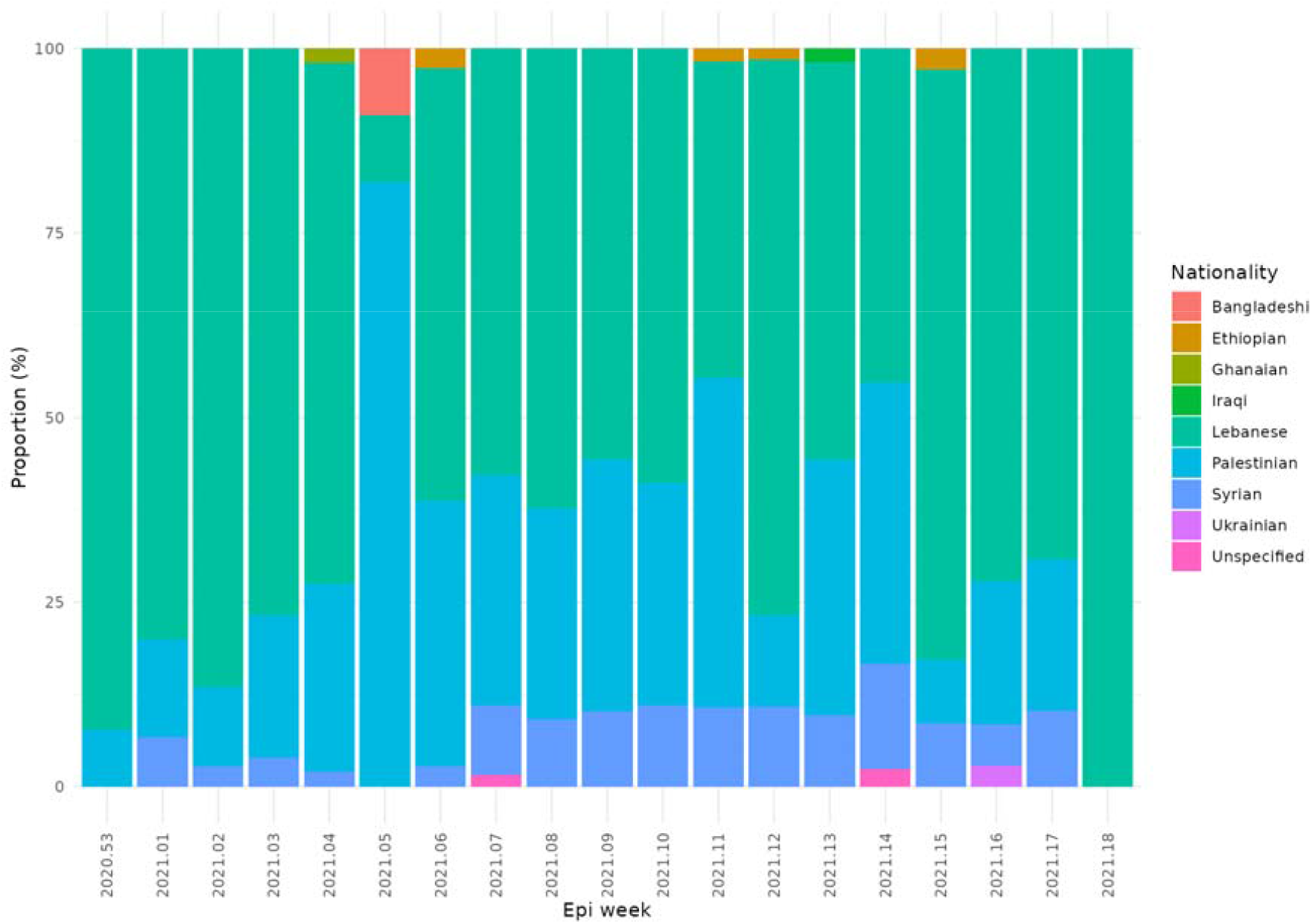
The proportion of self-declared nationalities of cases by week from January to May 2021.

Samples with a diagnostic polymerase chain reaction (PCR) cutoff value of less than 25 cycles were selected to maximise the chance of yielding a high quality genome. Of the 8 provinces, 2 (Beirut and Akkar) were underrepresented compared to their populations. Samples were selected with low CT levels, from different families and sites, whatever was the nationality.

The samples were collected from 29/12/2020 to 4/5/2021. Of these 96.68% (n=873 produced genomes of sufficient quality to allow for PANGO lineages to be called. Additionally, all samples (n=77) deposited in the GISAID SARS-CoV-2 public database and available on 2021-08-03 were added if they were noted as being from Lebanon, or from travellers from Lebanon. In particular, there were 20 additional sequences labelled Lebanon in GISAID that were known to be from travellers, which were removed from the analysis.

### Sentinel surveillance

The sentinel surveillance system for influenza and COVID-19 like illness was implemented in Lebanon in late 2020 with the objective to monitor circulation of influenza and SARS-CoV-2 viruses. The network includes 18 outpatient sites such as primary health care centers, hospital emergency departments, and clinics in camps for refugees and displaced populations. The sites cover 7 of the 8 Lebanese provinces. In each site, one day per week is dedicated for the surveillance where any patient fitting the WHO case definitions for Influenza or COVID-19 like illness, adapted from WHO, is included. On site, a trained team is responsible for data collection (variables related to identification, demographic, clinical and exposure) and sample collection (nasopharyngeal swab in mainly viral transport media/ viral stabilizer media). Clinical samples are referred on the same working day to the National Influenza Center (NIC) where RT-PCR is conducted for SARS-CoV-2. Positive samples are kept at NIC for potential genomic analysis. Positive cases are integrated into the national COVID-19 surveillance database.

### Field testing

COVID-19 field testing was initiated in Lebanon starting 2020. The objective is to provide free PCR testing for suspected cases and close contacts. The field sites are organized in partnership with municipalities, hospitals, MSF, UNHCR, UNRWA, and NGOs. All Lebanese districts are covered with at least 1 site per week. Target persons are: 1) COVID-19 suspected or probable cases of COVID-19, 2) close contacts adopting WHO case definitions, detected either via MOPH hotline or case investigation. At the site, a trained team proceeds with data collection and specimen collection (nasopharyngeal swab in viral transport media/viral stabilizer media). Samples are referred to designated laboratories, in particular the Laboratory of Molecular Biology and Cancer Immunology, Faculty of Sciences, Lebanese University (Hadath campus), where RT-PCR is conducted. Positive samples are also kept for potential genomic analysis. Positive cases are integrated into the national COVID-19 surveillance database.

### Genome sequencing and analysis

Samples were transported to the UK for high throughput genome sequencing. Primary samples were inactivated at the Department of Microbiology at the Norfolk and Norwich University Hospital Foundation Trust and extracted at the Bob Champion Research and Education laboratory at the University of East Anglia. Viral RNA was extracted using the Thermo Fisher Scientific MagMAX^™^ Viral/Pathogen II (MVP II) Nucleic Acid Isolation CE-IVD kit where 265 μl of Binding Buffer was added to 200 μl of the primary sample in a 96-deepwell plate and incubated at room temperature for 15 mins to inactivate the sample. Following sample inactivation 5 μl of Proteinase K and 10 μl of Binding Bead Mix were added. The remaining extraction steps were performed using the Thermo Scientific Flex System and the MVP_2wash_200_flex program using 500 μl of wash buffer, 500 μl 80% ethanol followed by elution of RNA in 50 μl molecular grade, RNAse free water. Viral RNA was converted in cDNA and was then amplified using the ARTIC protocol v3 (LoCost) (Quick 2020, 3) with sequencing libraries prepared using CoronaHiT (Baker et al. 2021). G genome sequencing was performed using the Illumina NextSeq 500 platform with one positive control and one negative control per 94 samples.

The raw reads were demultiplexed using bcl2fastq (v2.20). The reads were used to generate a consensus sequence using the ARTIC bioinformatic pipeline (*Ncov2019-Artic-Nf*, n.d.). Briefly, the reads had adapters trimmed with TrimGalore (Krueger [2016] 2020) and were aligned to the WuhanHu-1 reference genome (accession MN908947.3) using BWA-MEM (v0.7.17) (Li 2013); the ARTIC amplicons were trimmed and a consensus built using iVAR (v.1.3.0) (Grubaugh et al. 2019). PANGO lineages assigned using Pangolin (v3.1.7) and PangoLEARN model dated 2021-07-28 (Rambaut et al. 2020).

### Phylogenetic and clustering analysis

For the phylogenetic analysis, all sequences from GISAID where Lebanon is the country of exposure (Supplementary Table 2) were downloaded and added to the sequences from the current study. All remaining sequences from GISAID were then compared to this data set where we kept the closest ones for subsequent analysis to provide context. The alignment and neighbour search were done with uvaia (*https://github.com/quadram-institute-bioscience/uvaia*), and problematic (homoplasic or difficult to sequence) sites were masked from the alignment (Turakhia et al. 2020). Sequences with more than 50% N were excluded from analysis. For sequences with complete date information we estimated divergence times using treetime using an autocorrelated molecular clock. In particular, we used the autocorrelated clock model to estimate branch-wise substitution rates for all sequences, and then we removed the outlier sequences, from the 5% higher percentile of evolutionary rates. We estimated divergence times from the remaining samples assuming a mean clock rate of 8×10^−4^.

The number of transmission events into or outside Lebanon was found by ancestral reconstruction of nodes using castor for R, which calculates the probability of each ancestral node belonging to Lebanon or not based on the country of exposure information at the tips. Importation events are then the nodes with P (Lebanon) < 50% with at least one child node with P (Lebanon) = 100%, and exportation nodes are equivalently nodes with P (Lebanon)>50% with one or more nodes where P (Lebanon)=0%. The exposure information comes from the “Location” information on the GISAID metadata, and may contain errors (in case the data providers cannot ascertain the travel history of samples).

To find the closest matches per country, we used uvaia to compare all Alpha samples from Lebanon to the remaining sequences from GISAID, and keep the 128 closest neighbours per sample. Since the same country may be represented more than once amongst these 128 neighbours, the best match per country is the sample with the highest number of perfect matches for each country. Not all countries are represented in all samples, and a “perfect match” considers only the four bases ACGT (any other disagreement is a mismatch, where only sites excluding Ns are considered).

